# Face covering adherence is positively associated with better mental health and wellbeing: a longitudinal analysis of the CovidLife surveys

**DOI:** 10.1101/2020.12.18.20248477

**Authors:** Drew Altschul, Chloe Fawns-Ritchie, Alex Kwong, Louise Hartley, Clifford Nangle, Rachel Edwards, Rebecca Dawson, Christie Levein, Archie Campbell, Robin Flaig, Andrew McIntosh, Ian Deary, Riccardo Marioni, Caroline Hayward, Cathie Sudlow, Elaine Douglas, David Bell, David Porteous

## Abstract

Face masks or coverings are effective at reducing airborne infection rates, yet pandemic mitigation measures, including wearing face coverings, have been suggested to contribute to reductions in quality of life and poorer mental health. Longitudinal analyses of more than 11,000 participants across the UK found no association between lower adherence to face covering guidelines and poorer mental health. The opposite appears to be true. Even after controlling for behavioral, social, and psychological confounds, including measures of pre-pandemic mental health, individuals who wore face coverings “most of the time” or “always” had better mental health and wellbeing than those who did not. These results suggest that wearing face coverings more often will not negatively impact mental health.

## Introduction

Regulatory bodies and governments around the world recommend wearing face masks, termed ‘face coverings’ by the UK government, to control the spread of the 2019 novel coronavirus SARS-Co-V2 (Klompas et al., 2020) because face coverings are an effective low-cost measure for reducing the spread of infectious aerosols and droplets (Fischer et al., 2020). Wearing face coverings thus helps protect others from catching coronavirus, reducing spread (Lyu & Wehby, 2020), although high adherence to face covering guidelines is necessary for this to have an impact at the population level (Eikenberry et al., 2020).

Many of the most effective measures that reduce coronavirus transmission, such as distancing, have negative impacts on individual wellbeing and mental health at the population level (Qiu et al., 2020; Rossi et al., 2020). Since the pandemic began increases in loneliness, stress, anxiety, and depression, and decreases in life satisfaction and wellbeing have been reported (Kwong et al., 2020; Luchetti et al., 2020; Salari et al., 2020; Satici et al., 2020).

Wearing face coverings does not have obvious, direct links to negative experiences such as self-isolation or quarantine (Brooks et al., 2020), but might induce negative experiences through physical discomfort, communication difficulties, or stigmatization (Czypionka et al., 2020). This been the topic of public and informal debates (Czypionka et al., 2020; Howard et al., 2020), which often do not take evidence into account. The public confusion this debate creates may in turn drive non-compliance (Lyu & Wehby, 2020). Evidence for or against an impact of wearing face coverings on individuals’ lived experience would be valuable.

The CovidLife surveys^1^ are a longitudinal UK-wide study of over 18,000 individuals begun during the early stages of the 2020 lockdown. Here, we used CovidLife data across surveys to investigate the relationships between adherence to guidelines on face coverings and wellbeing, life satisfaction, anxiety, depression, and loneliness.

## Methods

### Data collection and study sample

UK residents aged 18 years and over were eligible to take part in the CovidLife surveys^2^. Data were collected via the Qualtrics platform. Data collection for Survey 1 commenced on 17 April and closed to new responses on 7 June 2020. This period overlapped with the first period of UK-wide ‘lockdown’. Survey 2 data were collected 21 July to 17 August 2020. This corresponded period when the UK government made face coverings mandatory on public transport and in many shops. More than 18,000 individuals responded to Survey 1, and of those that shared their email contact address, more than 11,000 returned to participate in Survey 2.

### Variables and data processing

All mental health outcome measures used here were asked in Survey 2 (Figure 1). In Survey 1, we asked individuals about their sense of loneliness and life satisfaction before and during lockdown. Mental health was assessed using common self-report instruments (e.g. Patient Health Questionnaire for depression – see below), which could be scored to create continuous outcomes (except loneliness, which was ordinal).

**Fig 1.**
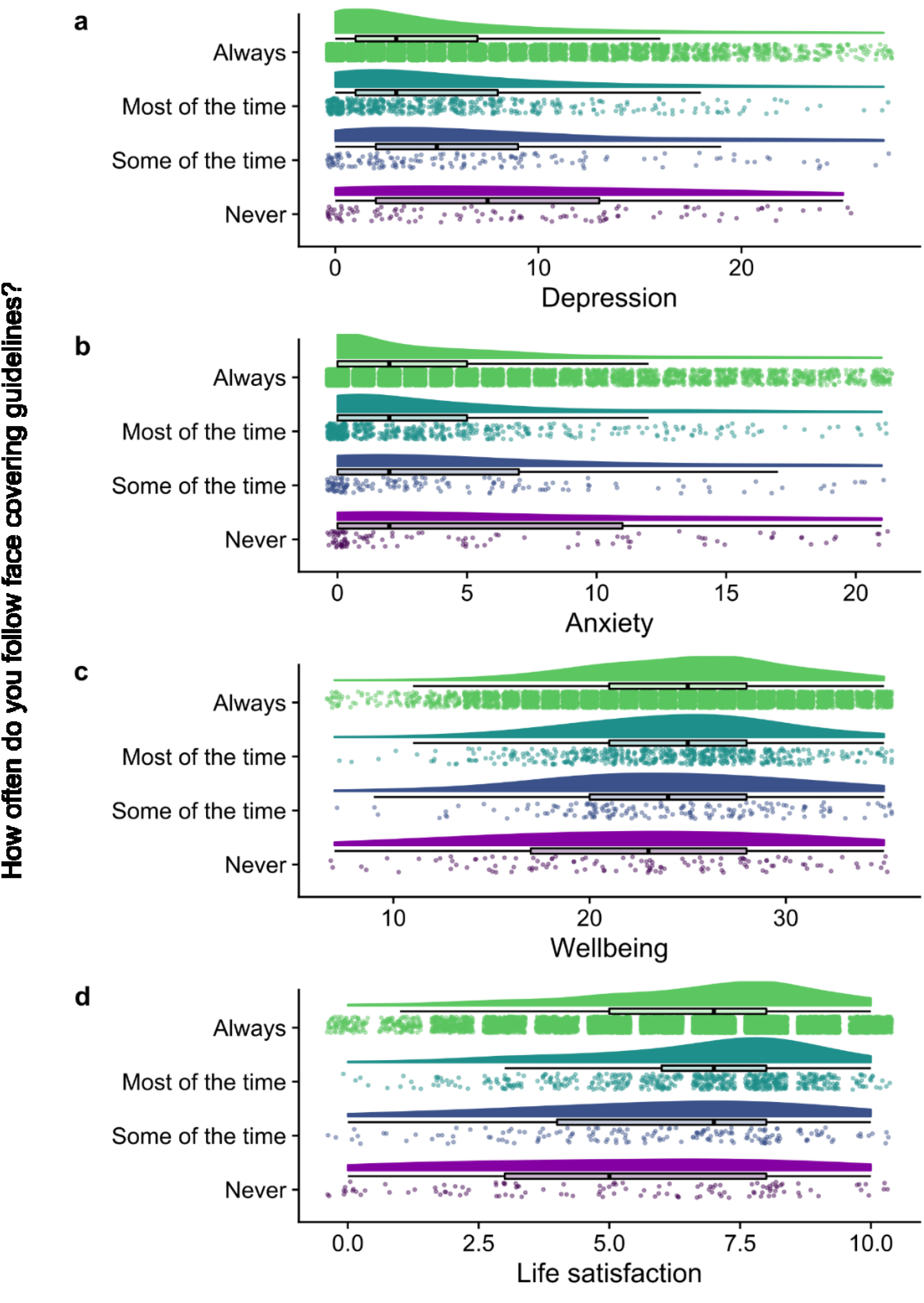
Raincloud plots of adherence to face coverings and main outcome variables wellbeing, life satisfaction, anxiety, and depression. The cloud portion of each plot is the smoothed distribution of all members of each category. The rain portions below show the raw, jittered observations that constitute the distributions. Boxplots illustrate the means, hinges represent the first and third quartiles, and the whiskers represent 1.5x the inter-quartile range. **a**. depression scored from 0 to 27, **b**. anxiety scored 0 to 21, **c**. subjective wellbeing scored from 7 to 35, **d**. life satisfaction scored from 0 to 10. Loneliness was not plotted in this manner due to the ordered nature of the data.

Unless otherwise stated, the variables with n ∼ 18,000 were collected during Survey 1 (e.g. pre-COVID-19 mental health) and variables with n ∼ 11,000 were collected during Survey 2 (e.g. adherence to face covering guidance).

### Depression

Depression was assessed with the Patient Health Questionnaire (PHQ-9) (Kroenke et al., 2001), which consisted of 9 questions asking about depressive symptoms. Each question was scored from 1 to 4, with higher values indicating increased frequency of symptoms. The PHQ-9 was administered in both Surveys 1 and 2. Sum scores were created which ranged from 0 to 27 (M = 4.53, SD = 5.20, n = 10,408 for scores assessed in Survey 2). Binary categorization used the recommended cut-off for possible depression (≥10).

### Anxiety

Anxiety was assessed with the Generalized Anxiety Disorder assessment (GAD-7) (Spitzer et al., 2006), which consisted of 7 questions asking about the presence of generalized anxiety disorder symptoms. Each question was scored from 1 to 4, with higher values indicating increased frequency of symptoms. The GAD-7 was administered in Surveys 1 and 2. Sum scores were created which ranged from 0 to 21 (M = 3.65, SD = 4.66, n = 10,608 for scores assessed in survey 2). Binary categorization used the recommended cut-off for possible anxiety (≥10).

### Wellbeing

Subjective psychological wellbeing was assessed with the Warwick-Edinburgh Mental Wellbeing Scale (WEMWBS) (Tennant et al., 2007), which consisted of 7 items. Each question was scored from 1 to 5, with higher values indicating better wellbeing. The WEMWBS was administered in surveys 1 and 2. Sum scores were created which ranged from 7 to 35 (M = 24.82, SD = 5.03, n = 11,084 for scores assessed in Survey 2). Binary categorization used the recommended cut-off (≤17).

### Loneliness

Loneliness was assessed with a single question asking “*How often have you felt lonely during the past week?*” (Altschul et al., 2020; Solano, 1980). Loneliness prior to lockdown was assessed with a similar question: “*Think back to before COVID-19 measures were introduced (i*.*e*., *January 2020), how often did you feel lonely then?*” Participants could choose between “*None, or almost none of the time*”, “*Some of the time*”, “*Most of the time*”, “*All, or almost all of the time*”, “*Don’t know*”, “*Prefer not to answer*” in response to both questions. For the period before COVID-19, 13,560 people (77%) reported being lonely “*almost none of the time*”, 3,781 (21%) were lonely “*some of the time*”, 240 (>1%) were lonely “*most of the time*” and 72 (<1%) were lonely “*all, or almost all the time*”. By the time of Survey 2, 7,957 people (77%) were lonely “*almost none of the time*”, 2,703 (24%) were lonely “*some of the time*”, 331 (3%) were lonely “*most of the time*” and 143 (>1%) were lonely “*all, or almost all the time*”. Note that the number of respondents in Survey 2 (n=17,653) was less than in Survey 1 (n=11,134). For the purposes of binary categorization, individuals who answered “*most of the time*”, or “*all, or almost all of the time*” were classified as being lonely, and others were not.

### Life satisfaction

Life satisfaction (Mazaheri & Theuns, 2009; Pavot et al., 1991) was assessed with a single question asking “*how satisfied are you with your life nowadays?*” Life satisfaction prior to the pandemic was assessed with the question “*Thinking back to just before the COVID-19 measures were introduced (i*.*e*., *January 2020), how satisfied were you with your life then?*” Participants were asked to answer the question using a 0 to 10 scale, where 0 indicated being not at all satisfied with life, and 10 indicated being extremely satisfied with life.

### Prior mental health diagnoses

In Survey 1, participants were categorized as having a mental diagnosis relevant to anxiety or depression if they reported being diagnosed with any of the following: “*Anxiety, nerves or generalised anxiety disorder*”, “*Depression*”, “*Mania, hypomania, bipolar or manic-depression*”, “*Panic attacks*”, or “*Social anxiety or social phobia*”. In Survey 1, 5,729 individuals had at least one such diagnosis, and 12,016 did not. 311 chose not to answer.

### Face covering

In a response matrix, participants were asked about various government guidelines: “*Have you been following the government guidance on*” and a list followed. The particular prompt under study was “*Wearing face coverings on public transport and in shops*”. Participants could respond “*Always*” (coded 4, n = 10,180, 92%), “*Most of the time*” (coded 3, n = 592, 5%), “*Some of the time*”, (coded 2, n = 172, 2%), and “*Never*” (coded 1, n = 120, 1%).

### Age & sex

Participants were asked their date of birth in Survey 1 and age was calculated from this (M = 56.6, SD = 14.34, n = 18,328). After this, participants were asked “*What is your sex? As assigned at birth*” and could answer “*Male*” (coded 1, n = 5999, 33%), “*Female*” (coded 2, n = 12,299, 67%), or “*Prefer not to answer*” (n = 125, <1%).

### Personality

30 questions from the 50-item International Personality Item Pool 5 factor instrument (Goldberg et al., 2006; Gow et al., 2005), those used to assess conscientiousness (M = 37.85, SD = 6.16, n = 17,356, 94% completed the questions), extraversion (M = 30.58, SD = 8.02, n = 17,424, 95% completed), and emotional stability (M = 33.56, SD = 8.45, n = 17,425, 95% completed), were asked during Survey 1.

### Psychological resilience

Resilience was measured using the Brief Resilience Scale (Smith et al., 2008), which consists of 6 questions rated from 1 (“*strongly agree*”) to 5 (“*strongly disagree*”). A sum score of these items was constructed to represent overall trait resilience (M = 21.34, SD = 4.94, n = 11,107, 98% completed).

### Living circumstances

In Survey 1, participants were asked “*Including yourself, how many people live in your household?*” and could answer anywhere between 1 and 12+ (M = 1.28, SD = 1.11, n = 17,955). In Survey 1, participants were asked “*What type of accommodation do you live in?*” and could choose from options: “*House or bungalow*”, “*Flat or apartment*”, “*Hostel*”, “*Mobile home or caravan*”, “*Sheltered housing*”, “*Homeless*”, “*Other*”, and “*Prefer not to answer*”. More affluent accommodations were lower in value (e.g. “*house*” was recorded as 1, the lowest value on the scale) (M = 1.23, SD = 0.48, n = 17,172). In Survey 1, participants were asked “*How many rooms are there in your house? Count living rooms, bedrooms, kitchens, utility rooms and studies. Do not count toilets, bathrooms, halls, landings, or cupboards*”. Participants could answer anywhere from 1 to 15+ (M = 6.01, SD = 2.06, n = 17,185). In Survey 2, participants were asked “*Do you have a partner that you live with? This could be someone you are married to/in a civil relationship with, or a person with whom you are co-habiting*”. Participants could answer “*Yes, I live with a partner*” (coded 1, n = 8,327), “*No, I do not live with a partner*” (coded 0, n = 2,864), and “*Prefer not to say*” (n = 311).

### Student status

Whether a participant reported being a student (coded 0, n = 17,592), a part-time student (coded 1, n = 314), or a full-time student (coded 2, n = 378).

### Self-rated health

Both general and mental health were assessed. Participants were asked “*In general, would you say your health is*” and “*In general, would you say your emotional or mental health is*” and could answer between “*excellent*” (1) and “*poor*” (5). General (M = 2.45, SD = 1.01, n = 18,307) and mental (M = 2.42, SD = 1.03, n = 18,305) health responses were comparable.

### Educational qualification

Participants were asked “*What is the highest educational qualification you have obtained?*” Responses available were “*Postgraduate degree*”, “*Undergraduate degree*”, “*Other professional or technical qualification*”, “*NVQ or HND or HNC or equivalent*”, “*Higher grade, A levels, AS levels or equivalent*”, “*Standard grade, National 4 or 5, O levels, GCSEs or equivalent*”, “*CSEs or equivalent*”, “*School leavers certificate*”, “*Other (please specify)*” with an attached open field to indicate the type of other, non-high school qualification, “*No qualifications*”, and “*Prefer not to answer*”. The scale ran from 1 to 10, with 1 representing “*No qualifications*” and 10 representing “*Postgraduate degree*” (M = 7.88, SD = 2.18, n = 17,059).

### Contact outside your household

In Survey 2, participants were asked “*When leaving your home, how likely are you to come into close contact with someone not living in your household? By close contact, we mean coming within 2 metres of someone*”. Participants could answer “*I don’t leave my home*” (1), “*Not at all likely*” (2), “*Not that likely*” (3), “*Somewhat likely*”

(4) or “*Very likely*” (5). M = 3.70, SD = 0.99, n = 11,267. Participants were also separately asked “*How regularly do you do these activities now?*” about the several social activities. The answers available were “*Every day/almost every day*” (6), “*3-4 days a week*” (5), “*1-2 days a week*” (4), “*Less than once a week*” (3), “*Rarely*” (2), and “*Never*” (1). The particular prompts relevant to the study at hand were “*meet*[ing] *with family members face-to-face*” (M = 3.32, SD = 1.46, n = 11,046) and “*meet*[ing] *with friends face-to-face*” (M = 2.88, SD = 1.19, n = 11044).

### Risk from getting COVID

In both Survey 1 and 2 participants were asked “*Do you think that you have had, or currently have COVID-19?*” Possible responses were “*Yes, confirmed by a positive test*” (n = 60, <1%), “*Yes, suspected COVID-19 but was not tested*” (n = 1,205, 11%), and “*No*” (n = 10,020, 89%). Participants were also asked “*Have you been contacted by letter or text message to say you are at severe risk from COVID-19 due to an underlying health condition and should be shielding?*” and could answer “*Yes*” (n = 1,423, 8%) or “*No*” (n = 16,881, 92%).

### Statistical analyses

Both linear and logistic regression models were used to investigate the associations between following guidance on wearing face coverings and measures of mental health and wellbeing. These models were longitudinal in that they allowed us to control for potential confounders including assessments of the outcomes (mental health and wellbeing) measured earlier, as well as age, sex, personality, living circumstances, education, resilience, physical health, and behavioral factors such as frequency of leaving one’s home, and meeting others (see section above). All analyses were conducted using the R programming language, version 3.6.1 (Ihaka & Gentleman, 1996). Analytic code is available on GitHub^3^.

### Ethical approval

The CovidLife study was reviewed and given a favorable opinion by the East of Scotland Research Ethics Committee (Reference: 20/ES/0021 AM02).

## Results

Mean mental health and wellbeing scores were lower for individuals who adhere to face covering guidance less often (Table 1), except for anxiety. Mental health among less adherent groups also appears to be more broadly distributed.

**Table 1.** Standardized regression coefficients and 95% confidence intervals for fully adjusted linear regression models of following government guidance on wearing face coverings and mental health and wellbeing outcomes.

| Variable | Outcomes |  |  |  |  |
| --- | --- | --- | --- | --- | --- |
|  | Depression<br>*ordinary least squares* | Anxiety<br>*ordinary least squares* | Loneliness<br>*ordered logistic* | Life satisfaction<br>*ordinary least squares* | Wellbeing<br>*ordinary least squares* |
| **Following guidance on wearing face coverings (higher values indicate more adherence)** | **-0.04\*\*\* (-0.05, -0.02)** | **-0.02\* (-0.04, -0.005)** | **-0.11\* (-0.21, -0.02)** | **0.04\*\*\* (0.02, 0.06)** | **0.02\*\* (0.01, 0.03)** |
| Age | -0.15*** (-0.17, -0.13) | -0.08*** (-0.11, -0.06) | 0.0001 (-0.13, 0.13) | 0.02 (-0.01, 0.04) | 0.06*** (0.04, 0.08) |
| Sex (female) | 0.04*** (0.03, 0.06) | 0.02 (-0.0001, 0.03) | 0.08 (-0.03, 0.18) | -0.05*** (-0.07, -0.03) | -0.01 (-0.02, 0.01) |
| Conscientiousness | -0.05*** (-0.06, -0.03) | 0.03*** (0.01, 0.05) | 0.11* (0.004, 0.21) | -0.01 (-0.02, 0.01) | 0.06*** (0.05, 0.08) |
| Extraversion | 0.01 (-0.005, 0.02) | 0.04*** (0.03, 0.06) | 0.34*** (0.24, 0.45) | -0.05*** (-0.06, -0.03) | 0.01 (-0.01, 0.02) |
| Emotional stability | -0.25*** (-0.27, -0.23) | -0.29*** (-0.31, -0.27) | -0.60*** (-0.74, -0.45) | 0.06*** (0.04, 0.08) | 0.19*** (0.17, 0.21) |
| Number of people living with | -0.004 (-0.02, 0.01) | 0.04*** (0.02, 0.06) | -0.07 (-0.20, 0.06) | -0.01 (-0.03, 0.01) | -0.02* (-0.04, -0.003) |
| Accommodation type | 0.02* (0.004, 0.04) | 0.02* (0.005, 0.04) | 0.07 (-0.04, 0.17) | -0.03*** (-0.05, -0.01) | -0.03*** (-0.04, -0.01) |
| Rooms in house | -0.01 (-0.03, 0.005) | -0.01 (-0.03, 0.004) | -0.18** (-0.30, -0.05) | -0.01 (-0.02, 0.01) | 0.01 (-0.01, 0.02) |
| Student (yes) | 0.04*** (0.02, 0.06) | 0.04*** (0.02, 0.06) | 0.09 (-0.02, 0.20) | -0.003 (-0.02, 0.02) | 0.01 (-0.01, 0.02) |
| General health | 0.10*** (0.08, 0.12) | 0.02 (-0.004, 0.04) | -0.02 (-0.15, 0.10) | -0.03*** (-0.05, -0.01) | -0.05*** (-0.06, -0.03) |
| Mental health | 0.22*** (0.19, 0.24) | 0.15*** (0.12, 0.17) | 0.68*** (0.53, 0.83) | -0.11*** (-0.13, -0.08) | -0.16*** (-0.18, -0.13) |
| Do you live with a partner? (yes) | -0.06*** (-0.07, -0.04) | -0.04*** (-0.06, -0.02) | -0.92*** (-1.03, -0.82) | 0.07*** (0.05, 0.09) | 0.08*** (0.06, 0.09) |
| Resilience | -0.11*** (-0.12, -0.09) | -0.21*** (-0.23, -0.19) | -0.74*** (-0.87, -0.61) | 0.24*** (0.22, 0.26) | 0.34*** (0.32, 0.36) |
| Educational qualification | 0.003 (-0.01, 0.02) | -0.001 (-0.02, 0.02) | -0.13* (-0.23, -0.03) | -0.02** (-0.04, -0.01) | -0.03*** (-0.05, -0.02) |
| Loneliness before lockdown |  |  | 1.14*** (1.04, 1.24) |  |  |
| Life satisfaction before lockdown |  |  |  | 0.27*** (0.25, 0.29) |  |
| Has an anxiety or depression diagnosis (yes) | 0.05*** (0.03, 0.07) | 0.04*** (0.02, 0.06) |  |  |  |
| How regularly do you meet with family? | 0.001 (-0.01, 0.02) | -0.01 (-0.02, 0.01) | -0.24*** (-0.34, -0.13) | 0.04*** (0.02, 0.05) | 0.06*** (0.04, 0.07) |
| How regularly do you meet with friends? | -0.04*** (-0.05, -0.03) | -0.07*** (-0.09, -0.05) | -0.24*** (-0.35, -0.14) | 0.06*** (0.04, 0.07) | 0.05*** (0.03, 0.06) |
| When leaving your home, how likely are you to come into close contact with others? | -0.003 (-0.02, 0.01) | -0.001 (-0.02, 0.01) | -0.23*** (-0.34, -0.13) | 0.01 (-0.01, 0.03) | 0.02* (0.004, 0.03) |
| Have you had COVID? (yes) | 0.04*** (0.02, 0.05) | 0.04*** (0.02, 0.05) | 0.25*** (0.16, 0.34) | -0.03*** (-0.04, -0.01) | -0.02** (-0.03, -0.01) |
| Are you at severe risk from COVID? (yes) | 0.01 (-0.01, 0.02) | 0.02** (0.01, 0.04) | 0.25*** (0.15, 0.35) | -0.03*** (-0.05, -0.02) | -0.01 (-0.03, 0.003) |
| Intercept | 0.01*** (0.004, 0.02) | 0.03*** (0.02, 0.04) |  | -0.01*** (-0.02, -0.01) | -0.02*** (-0.03, -0.01) |
| Observations | 9,544 | 9,745 | 10,268 | 10,370 | 10,373 |
| R<sup>2</sup> | 0.48 | 0.40 |  | 0.34 | 0.48 |
| Adjusted R<sup>2</sup> | 0.48 | 0.40 |  | 0.34 | 0.47 |
| Residual Std. Error | 0.34 (df = 9522) | 0.38 (df = 9723) |  | 0.40 (df = 10348) | 0.36 (df = 10352) |
| F Statistic | 415.70*** (df = 21; 9522) | 314.61*** (df = 21; 9723) |  | 259.31*** (df = 21; 10348) | 468.47*** (df = 20; 10352) |

Note:
\*p<0.05; \*\*p<0.01; \*\*\*p<0.005

Linear (and ordinal for loneliness) regression models of mental health outcomes are presented in Table 1. These models were fully adjusted for pre-COVID-19 mental health, which was operationalized through depression or anxiety relevant diagnoses for post-COVID-19 depression and anxiety scores, and through self-reported assessment of pre-COVID-19 loneliness and life satisfaction for those outcomes. Only subjective psychological wellbeing could not be adjusted in this way. Many other potential confounders were included (see the methods and Table 1). Better adherence to guidance on wearing face coverings was significantly associated with better mental health and wellbeing across all measures, even controlling for prior mental health and wellbeing, as well as other potentially confounding covariates.

The outcomes can also categorize individuals as having either poor mental health in a particular domain or not (Kwong et al., 2020). We fit logistic regression models with these outcomes, which give the odds of having poor mental health or wellbeing depending on degree of adherence. For each outcome, we fit basic and fully adjusted models. Basic logistic regression models controlled for age and sex. Fully adjusted models controlled for all the variables described in the methods section, as with our linear regression models. The results of the logistic regression models accord well with those presented in Table 1 and Figure 1. Except for depression, there were significant associations between wearing a face covering “*most of the time*” or “*always*” and better mental health. Odds ratio are illustrated in Figure 2, and the fully adjusted odds ratios are described in the following paragraphs.

**Fig 2.**
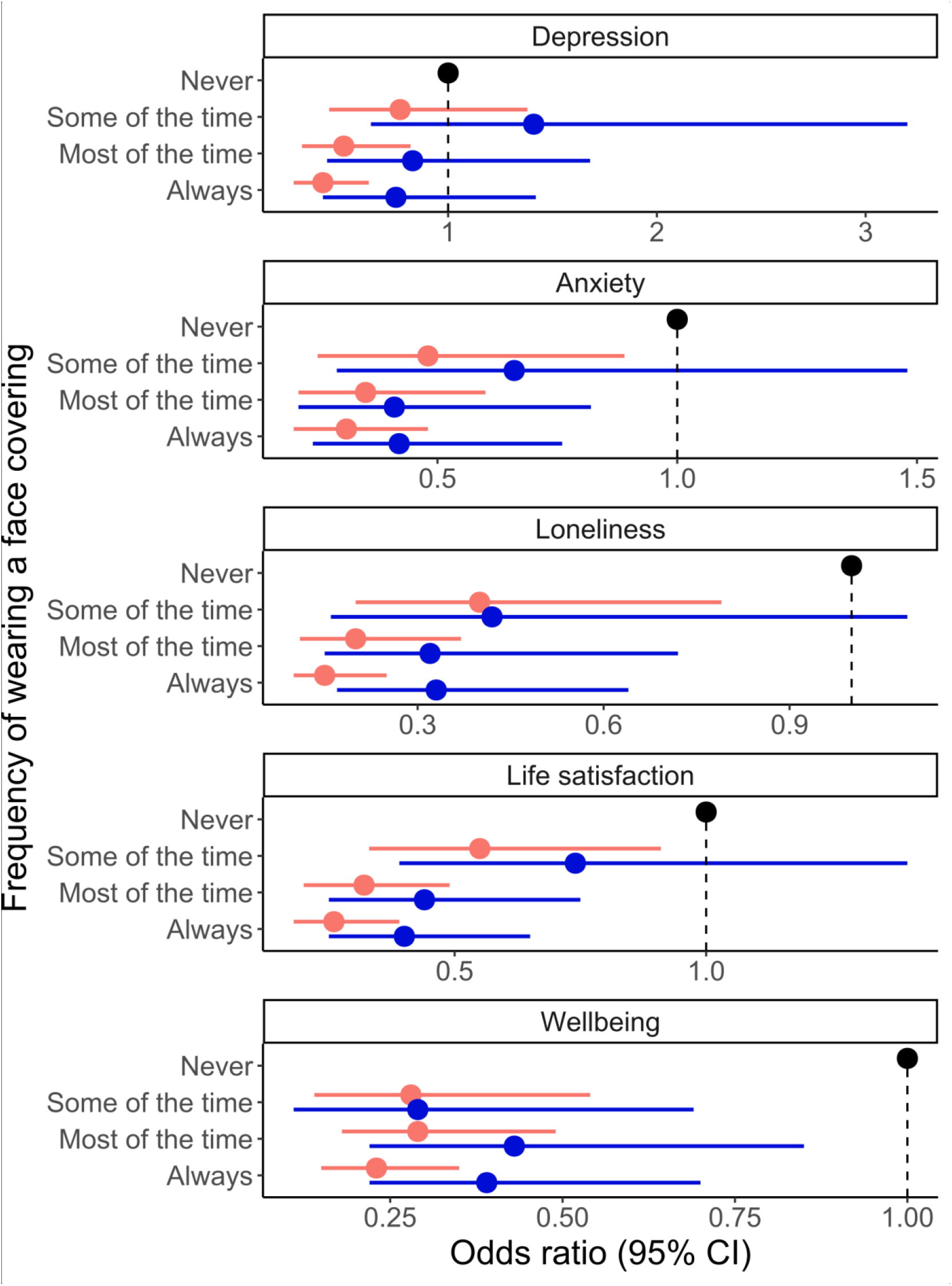
Minimally and fully adjusted odds ratios and confidence intervals for associations between wearing face coverings and mental health or wellbeing. All outcomes were categorized such that a positive event represented having poor mental health for that measure, so odds ratios (ORs) less than 1 indicate lower odds of having poor mental health or wellbeing. Light red dots and bars indicate ORs and 95% CIs for models adjusted for only age and sex, whereas blue dots and bars indicate ORs and 95% CIs for models fully adjusted for all relevant covariates (see methods and Table 1 for listings of covariates). Black dots indicate the reference category, which was always “*never*”.

The odds of feeling anxious were 58% lower among individuals who “*always*” adhered to guidance on wearing face coverings (adjusted OR=0.42, 95% CI=0.24 to 0.76, p=0.004), whilst the odds of having depressive symptoms were 25% lower among individuals who “*always*” adhered to guidance on face coverings (adjusted OR=0.75, 95% CI=0.40 to 1.42, p=0.36). The odds of feeling lonely most or all of time were 67% lower among individuals who always wore face coverings (adjusted OR=0.33, 95% CI=0.17 to 0.64, p<0.001), the odds of being satisfied with life were 60% higher among individuals who “*always*” wore face coverings (adjusted OR=0.40, 95% CI=0.25 to 0.65, p<0.001), and the odds of low wellbeing were 62% lower among individuals who “*always*” wore face coverings (adjusted OR=0.38, 95% CI=0.21 to 0.71, p=0.001).

Wearing a face covering “*some of the time*” was associated with 74% lower odds of poor wellbeing compared to those who “*never*” adhered (OR=0.26, 95% CI=0.10 – 0.67, p=0.006), but otherwise, wearing a face covering only “*some of the time*” was not significantly associated with good mental health. Although adhering to guidance on wearing face coverings “*most of the time*” was significantly associated with good mental health and wellbeing for all the same outcomes as “*always*” adhering, the associations were not as strong, except for loneliness and anxiety. Wearing face coverings “*most of the time*” appeared to have a slightly stronger association with less loneliness (OR=0.32, 95% CI=0.15 – 0.72, p=0.005) and anxiety (OR= 0.41, 95% CI=0.21 – 0.82, p=0.011).

## Discussion

Adhering to government guidance on wearing face coverings was not associated with poorer mental health or wellbeing, nor with a negative impact on mental wellbeing, all else being equal. Indeed, the opposite appears to be the case: stronger adherence to guidelines is associated with less anxiety and loneliness, and higher life satisfaction and wellbeing. Moreover, the relationships among wearing face coverings and having better mental health and wellbeing could not be explained by relevant psychological, medical, sociodemographic, or behavioral factors.

Many of our control variables were associated with multiple aspects of mental health and wellbeing (Table 1), yet the associations between wearing face coverings and mental health outcomes survived adjustment. For instance, trait extraversion, a measure of an individual’s overall sociality, as well as frequency of leaving one’s home for personal encounters or how often one meets with friends or family, are all indicators of how often an individual is likely to leave their home to interact with people. Close (unshielded) person-to-person interaction is far and away the most common way the coronavirus is spread (Desai & Patel, 2020; World Health Organization, 2020); it has a strong bearing on international guidance on the value of face coverings as an easily adopted, low cost mitigation measure. Nevertheless, including socialization variables as controls did not remove the association between wearing face coverings and mental wellbeing, suggesting that what a person does while wearing a face covering cannot be wholly responsible for mental wellbeing differences. Similarly, whether an individual already had poor mental health or wellbeing, or was predisposed to having poor mental wellbeing – either through low psychological resilience or having a previous mental health diagnosis – did not eliminate the association between wearing a face covering and better mental wellbeing. All this holds true for the range of socioeconomic and demographic variables we included that are known to relate to mental health and wellbeing outcomes (Stewart-Brown et al., 2015; Yu & Williams, 1999). There may simply be something about wearing a face covering that makes people feel safer, and reassured that they are “doing the right thing” for themselves and their community.

Relationships with wearing face coverings were found across all mental health and wellbeing measures, thus implying an underlying commonality. The only measure that was not fully consistent was depression, which may be due to relatively fewer reports of increased depressive symptoms post-pandemic, compared to other mental health and wellbeing measures (Kwong et al., 2020).

This study accords with earlier work that found that not adhering to guidance on wearing face coverings can be viewed negatively by others (Betsch et al., 2020). It reveals another side to adherence behavior: regardless of whether stigmatization or discomfort felt while wearing a face covering do or do not harm mental health and wellbeing, people who do not wear face coverings have lower mental health than those who do. Again, our results cannot be entirely explained by prior mental health or other factors.

Wearing face coverings in public can protect others from contracting coronavirus infections (Howard et al., 2020), but high uptake is necessary to prevent deaths from COVID (Eikenberry et al., 2020) and reduce stigma (Betsch et al., 2020), and voluntary policy does not appear to meet these thresholds (Eikenberry et al., 2020). Our findings from the CovidLife Surveys countermand speculation that face coverings may have a negative effect on mental health and wellbeing. Our data in fact provide strong evidence that following government guidance on face coverings is associated with better rather than poorer mental health and wellbeing. This evidence could be an important motivator for continued advocacy by policy makers and adherence by members of the public.

## Data Availability

Data used in this publication are available to bona fide researchers upon request to the Generation Scotland access panel via a standard application procedure.

## Footnotes

1 https://www.ed.ac.uk/generation-scotland/covidlife-volunteers/what-is-covidlife

2 https://www.ed.ac.uk/files/atoms/files/2020-05-15_covidlifesurvey_report_final_web.pdf

3 https://github.com/dmaltschul/FaceCovering_CovidLife

